# Evaluation of the effect of auto-transplanted tooth for repairing maxillary sinus perforation compared to traditional flap repair

**DOI:** 10.1101/2024.08.07.24311624

**Authors:** Fenglin Liao, Biao Zhang, Haoyan Zhong

## Abstract

**Objective:** The purpose of this study is to compare the outcomes of using auto-transplanted tooth and flap for repairing sinus perforation after tooth extraction. The aim is to provide a new reference for selecting appropriate repair methods for maxillary sinus perforation.

**Methods and materials:** This study involved 19 patients with sinus perforation who underwent treatment at the Department of Oral Surgery, Hospital of Stomatology Wuhan University, from March 2021 to September 2022. The study included two groups: a test group with 11 cases where perforation of the maxillary sinus mucosa greater than 5 mm was found during autologous tooth transplantation, and a control group with 8 cases where maxillary sinus mucosa perforation greater than 5 mm was found during tooth extraction. Clinical examination and radiographic examination were taken at 2 weeks, 1, 3, 6, and 12 months post-surgery of two groups. The two-year survival probability of the AT group was assessed using Kaplan-Meier survival analysis.

**Results:** The success rate of the auto-transplantation (AT) group was 72.7% (8/11), while the success rate of the flap transfer (FT) group was 100%. Upon analysis, it was found that the survival probability of AT group was significantly lower when the gingival index(GI) score was 2 (51.8%), in comparison to when it was 0 or 1 (100%).

**Conclusion:** The study findings demonstrated that the utilization of auto-transplanted teeth yielded a favorable outcome in restoring maxillary sinus perforation, suggesting its viability as a potential option.

## 1 INTRODUCTION

The maxillary sinus is a cone-shaped cavity located in the maxillary bone. It is lined with mucosa and surrounded by bone walls[1, 2]. Blood vessels and nerves that supply the teeth and periodontal tissue pass through either the intraosseous alveolar canal or beneath the mucosa[3, 4]. Maxillary sinus perforation refers to the formation of a new passage connecting the oral cavity and the maxillary sinus cavity[5, 6]. This condition often occurs due to tissue defects in the soft and hard tissues caused by procedures like tooth extraction. Studies suggest that factors such as smoking habits, sinus contour, and sinus membrane thickness are linked to maxillary sinus perforation[5, 7, 8]. There is controversy surrounding repair methods for perforations larger than 5mm[9]. Traditional repair methods include using removable plates for protection, surgical flaps (such as buccal mucosteal flap slide, palatal mucosteal flap transfer, and buccal fat pad transplantation), or autologous/non-autologous bone grafting[10, 11]. The local soft tissue flap method involves transferring soft tissue flaps around the alveolar fossa to promote healing of the perforated site. However, this repair plan requires highly skilled surgeons to ensure adequate blood supply to the transferred flap and buccal fat pad, as well as maintain good sealing ability of the soft tissue to the bone graft[12, 13]. Additionally, it increases surgical difficulty and may result in postoperative trauma and inability to restore the original gum shape and structure[12]. A systematic review and meta-regression analysis indicate that intraoperative sinus perforation can increase the risk of implant failure after sinus lift surgery[14]. However, a 6- to 20-year retrospective study found that neither sinus membrane perforation nor the presence of voids significantly affected implant survival as long as proper supportive maintenance therapy with good oral hygiene was provided[15].

Two case reports have shown that auto-transplantation of teeth can be an effective method for treating single defects with sinus perforation[16, 17]. Long-term observations over a period of 30 months revealed alleviation of inflammatory processes in the maxillary sinus area, along with restoration of the cortical plate. Satisfactory healing was also observed in the transplanted tooth area, with recovery of alveolar bone attachment. Based on these case reports, autologous tooth transplantation holds promise as a potential solution to the aforementioned problems. The digital design of autologous tooth transplantation technology has shown benefits in reducing periodontal membrane injury, improving the success rate of tooth transplantation, achieving good soft tissue morphology, and maintaining bone mass[18, 19]. Furthermore, the osteogenic potential of the periodontal membrane and maxillary sinus mucoperiosteum provide a theoretical basis for these cases[20, 21].

In light of these findings, we conducted a retrospective study to compare the outcomes of auto-transplanted tooth and flap in repairing sinus perforation after tooth extraction. We also assessed risk factors that may affect maxillary sinus membrane perforation and the effect of oral hygiene status on the restoration of maxillary sinus perforation with auto-transplanted tooth. The study compared clinical and imaging results one year after performing two different surgical methods. The choice of surgical method was based on the patient’s preferences and natural conditions, and a third molar without occlusal teeth was used as the donor tooth. This study aims to offer novel insights and ideas for the treatment plan of patients with maxillary sinus mucosa perforation.

## 2 METHODS

This study included 19 patients with sinus perforation who received treatment at the Department of Oral Surgery at the Hospital of Stomatology Wuhan University between March 2021 and September 2022. The study was approved by the Institutional Review Board of the Hospital of Stomatology Wuhan University {Approval number:[2021]伦审字 (B39)}. The cases were divided into an test group and a control group. The test group (11 cases) consisted of cases where perforation of the maxillary sinus mucosa greater than 5 mm was found during autologous tooth transplantation. The control group (8 cases) consisted of cases where maxillary sinus mucosa perforation greater than 5 mm was found during tooth extraction. In the test group, auto-transplanted tooth and concentrated growth factor (CGF)[Medifuge 200, Silfradent s.r.l., Italy] were used to repair maxillary sinus perforation, while in the control group, flap and collagen sponge(Type I Collagen,1.4*2.4cm)[Beiling, Beijing Yierkang Bioengineering Co., Ltd, China] were used for repair. Both groups of cases were performed by two experienced doctors. Patients with active infections or a history of head and neck radiotherapy, as well as those with other conditions that might affect wound healing, were excluded from the study. Additionally, individuals who declined to sign the informed consent or provide relevant documentation were also excluded. Demographic information, smoking history, apical shadow of affected tooth before surgery and oral hygiene status of the patients were collected. All patients underwent X-ray examination before and after surgery. The periapical radiograph (PA,eXpert DC Pennsylvania USA) was taken and images were assessed using Digora (Soredex, Helsinki,Finland). The cone-beam computed tomography (CBCT) was taken by the same machine (Newtom VGI, Quantitative Radiology, Verona, Italy) in all included cases. The CBCT parameters remained consistent throughout the study. The images were saved in DICOM 3.0 format, and the CT machine had a resolution of 0.30 mm. Image analysis and processing were performed using New-TomNNT software. CBCT measurements were taken after surgery to assess maxillary sinus mucosa lesion, perforation size, sinus membrane thickness, and alveolar bone mass. All patients provided informed consent. Figure 1 depicts the CT scans taken before, immediately after, and one year after the repair of the perforation of the maxillary sinus with transplanted teeth. Similarly, Figure 2 illustrates the CT scans captured before, three months after, one year after the perforation of the maxillary sinus using the flap technique.

**Figure 1.**
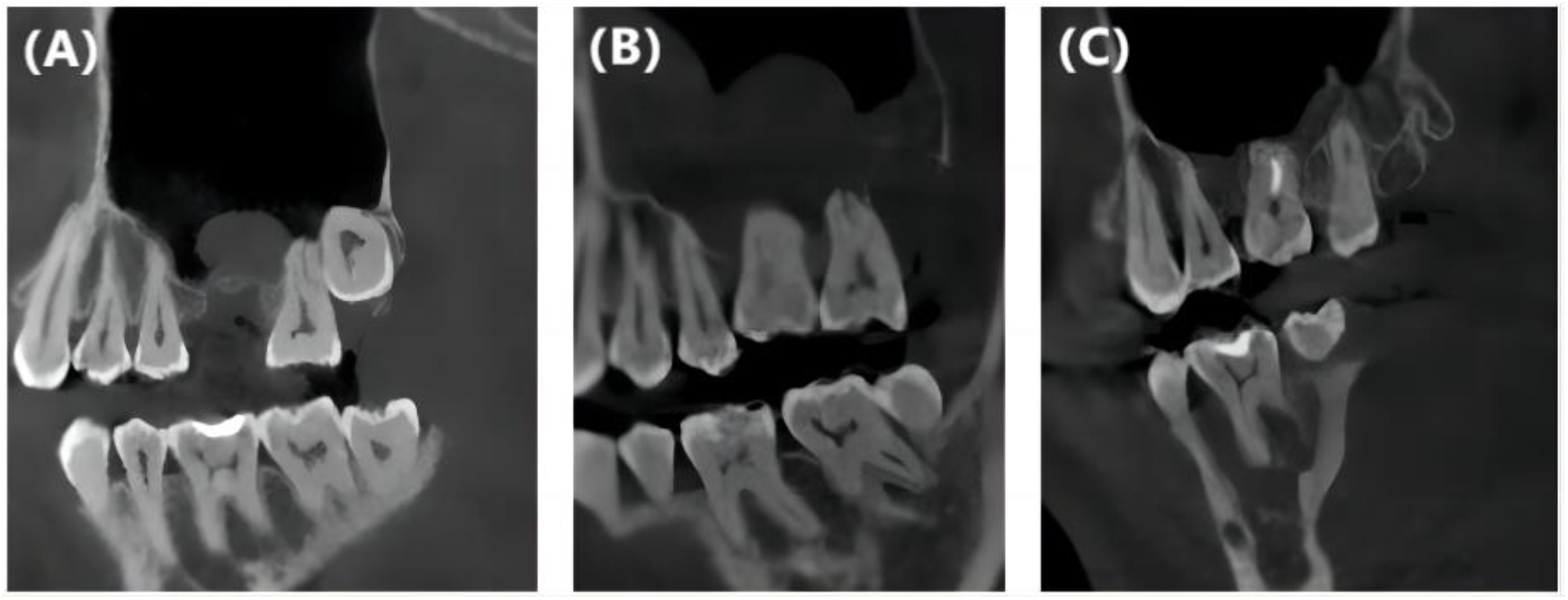
depicts the CT scans taken before(A), immediately after(B), and one year after the repair(C) of the perforation of the maxillary sinus with transplanted teeth.

**Figure 2.**
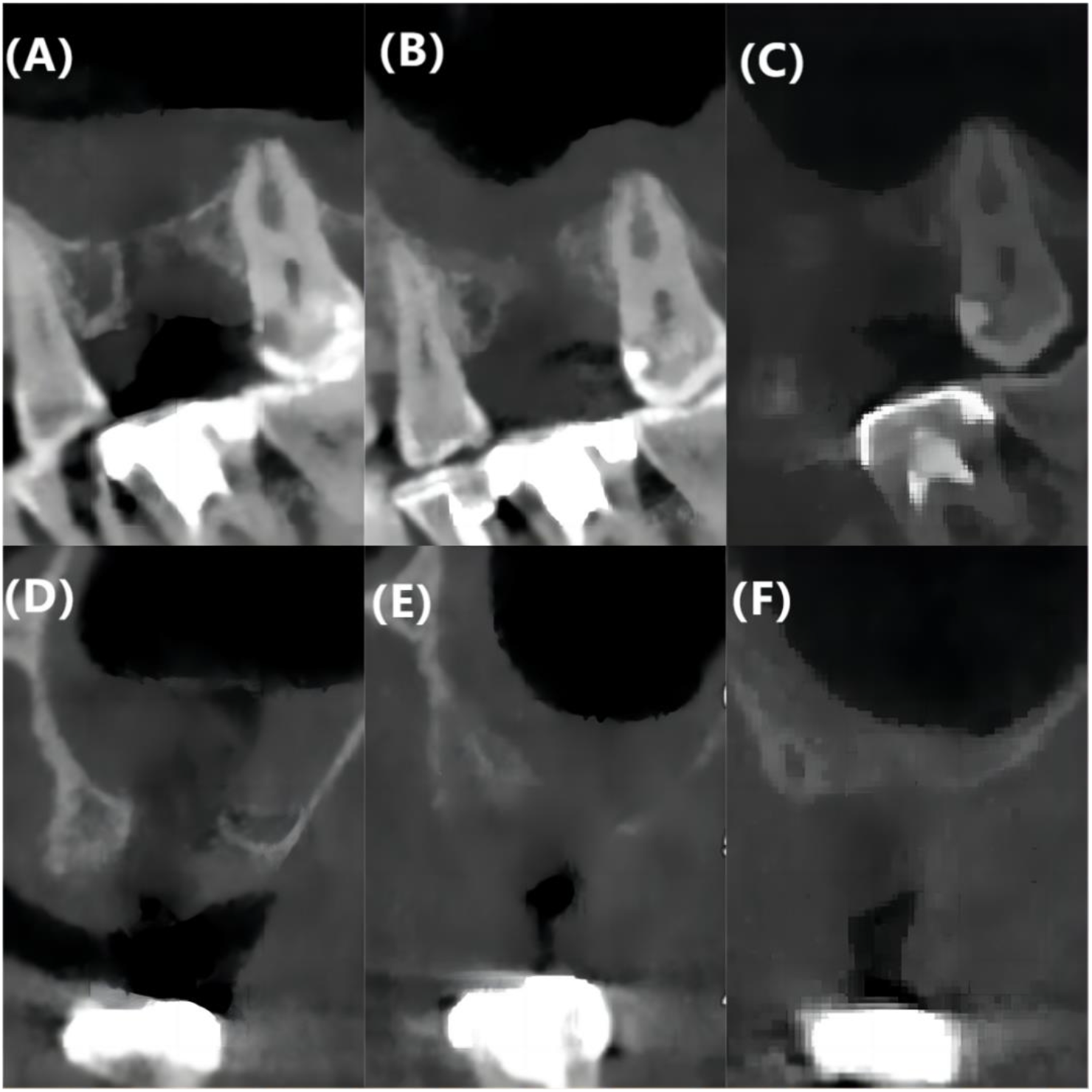
depicts the CT scans taken before(A,D), three months after(B,E), and one year(C,F) after the repair of the perforation of the maxillary sinus with tissue flap.

### Surgical procedure for the test group(Figure 3)

The optimal position of the donor tooth was determined using software simulation prior to surgery. An implant guide plate was designed using 3-matic software(3-matic 13.0, Materialize, Leuven, Belgium), which recorded the simulated implant position. Subsequently, the donor model and the implant guide plate were created using 3D printing technology. Under local anesthesia, an incision was made in the oral mucosa to completely remove the donor tooth. The maxillary sinus was internally lifted using the implant kit(Osstem Implant Co., Ltd., Seoul, Korea Korean). During the procedure, the sinus mucosa was perforated and a submucosal cyst in the maxillary sinus was removed. The recipient site was prepared using a model tooth, and the guide plate was used to confirm the orientation and depth of the implantation. CGF was placed at the site of the sinus floor perforation, followed by the implantation of the donor tooth, which was confirmed using the guide plate. gingival flap reduction and suturing were performed. Finally, an elastic ligature wire and resin were used to secure the donor tooth, with the bite separation using glass ionomer.

**Fig. 3.**
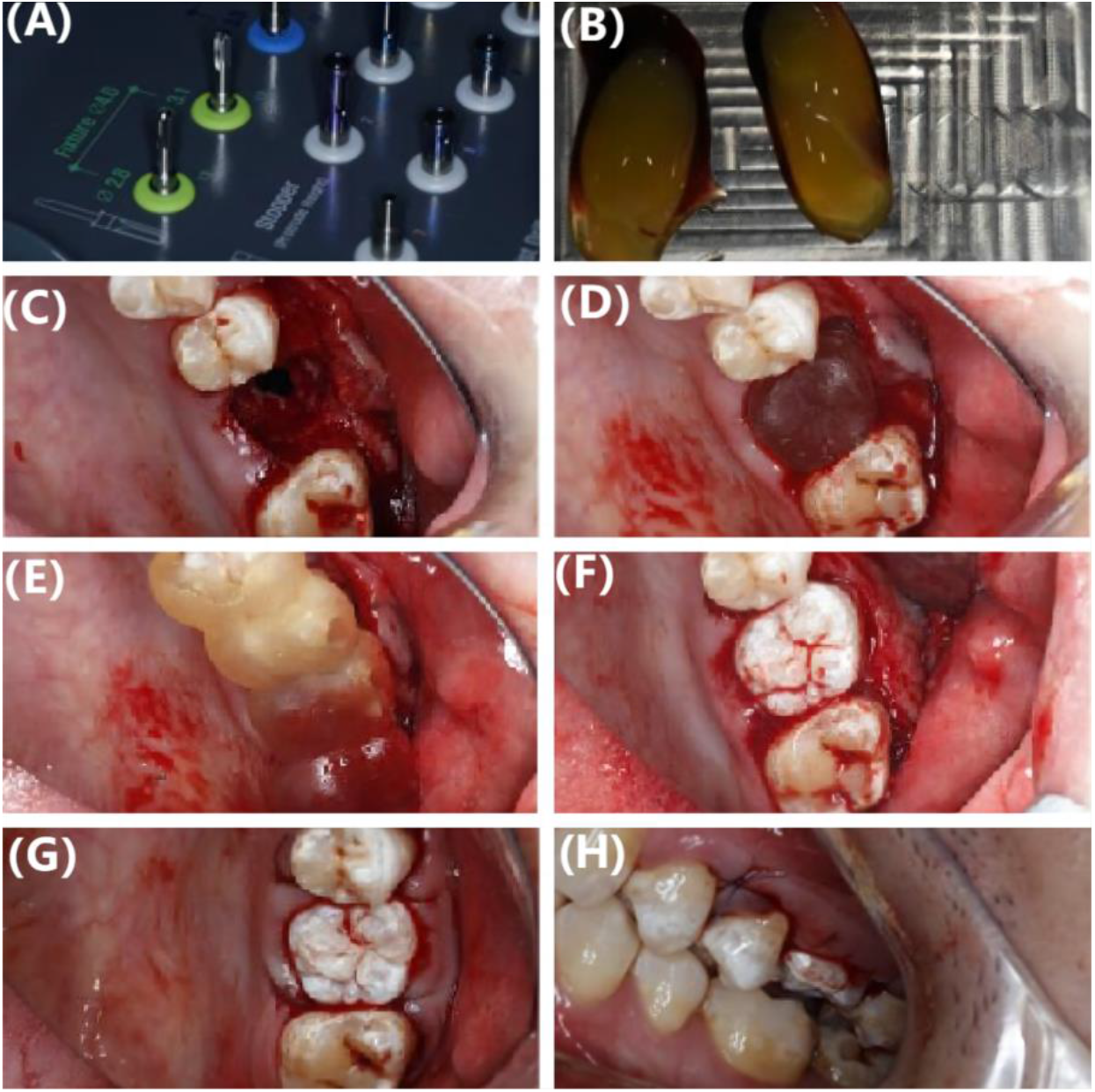
OSSTEM(KOREAN) internal lifting tool box(A),Intraoperative perforation of sinus floor mucosa(C),The model tooth was used to make the recipient socket(D),The orientation and depth of the transplanted tooth were confirmed by the guide plate(E),CGF was placed at the perforation of the sinus floor, and the donor tooth was confirmed to be in place with a guide plate(B,F). gingival flap reduction and suture(G,H).

### Surgical procedure for the control group(Figure 4)

Under local anesthesia, extract the affected tooth, clean the alveolar socket, the tooth extraction wound area was filled with a collagen sponge. A full-thickness mucosal flap was prepared on the palatal side of the tooth extraction wound to ensure that the base of the full-thickness mucosal flap was wide and completely contained the palatal neurovascular bundle, and then the mucosal flap was rotated across the tooth extraction wound and sutured to the buccal mucosa. Iodoform gauze was used to bandage the area of the palatal mucosal defect, followed by suturing the wound and applying compression to stop bleeding.

**Fig. 4.**
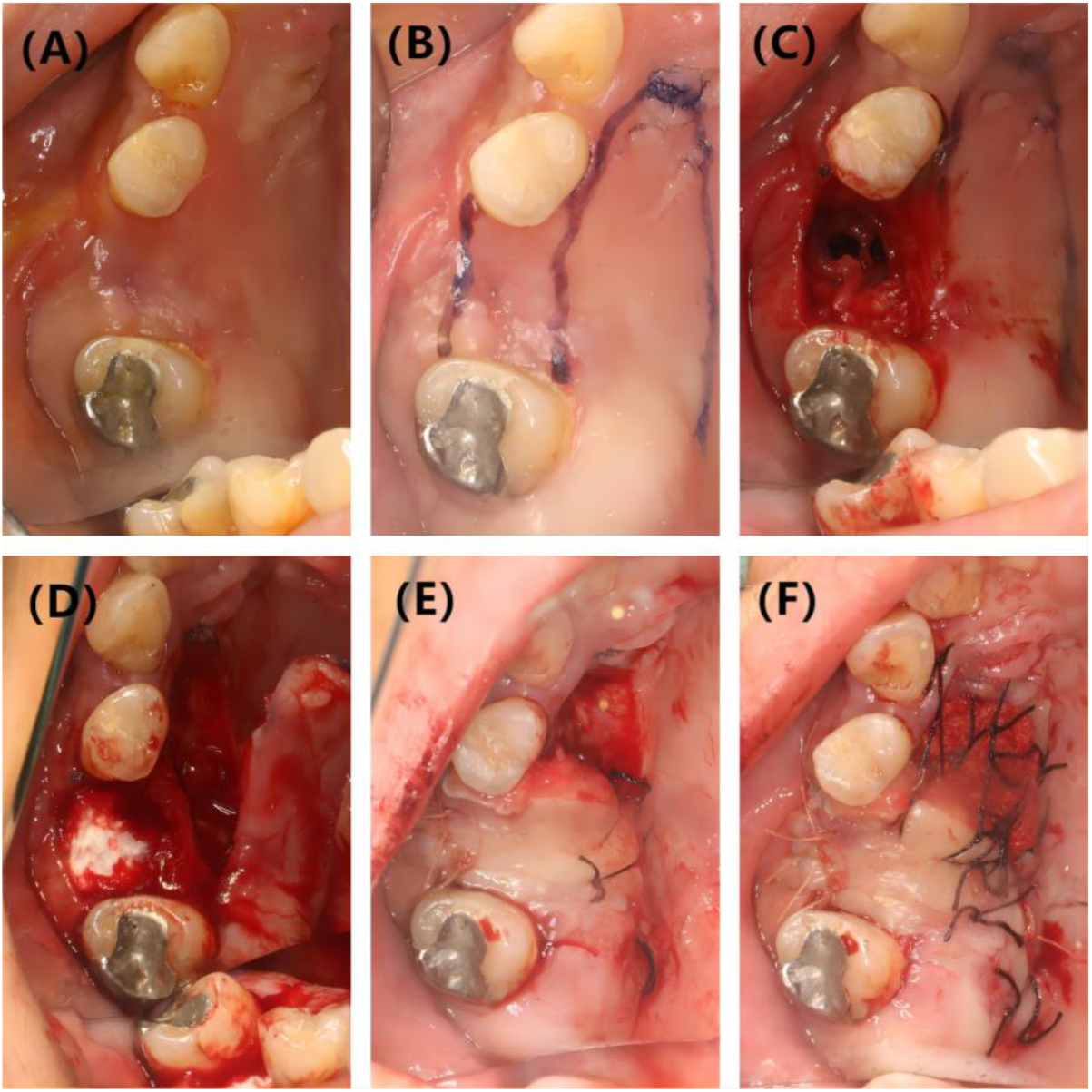
The preoperative photo of the operation area (A). The preoperative incision design (B). The maxillary sinus perforation (C). The tooth extraction wound area was filled with a collagen sponge (D). To repair the maxillary sinus perforation, an anterior palatine neurovascular pedicle flap was transferred from the palatal side (E). Iodoform gauze was then used to compress the palatal mucosal defect area, and the wound was sutured (F).

The recommended antibiotic regimen was 500 mg of Amoxicillin three times a day, starting 2 hours before surgery and continuing for 7 days after surgery. Patients were advised to take painkillers as needed. Additionally, Chlorhexidine (0.12%) mouthwash was recommended twice a day for 14 days. General postoperative instructions included avoiding blowing or sneezing for four weeks and refraining from chewing with the auto-transplanted tooth for eight weeks. Sutures were typically removed 10-14 days after surgery, and root canal treatment for the test group was completed 2-3 weeks after surgery. The fixator was removed during the follow-up visit after 4 weeks.

Clinical examination included recording percussion, mobility of transplanted tooth, and soft tissue healing at 2 weeks, 1, 3, 6, and 12 months post-surgery. gingival index (GI) was employed to assess the patients’ oral hygiene status. Imaging examinations were conducted as part of the study protocol. This included routine CBCT before the operation, as well as PA at two weeks and one month after the operation. CBCT was further utilized at 3, 6, and 12 months post-operation to observe the periapical bone formation of autografted teeth and the healing process of maxillary sinus perforation. Failure was defined as any sinus requiring secondary debridement or exhibiting poor clinical and radiographic manifestations of transplanted teeth.

The relevant statistical analysis was performed using SPSS software (SPSS/25.0, SPSS, Chicago, IL, USA).

## 3 Results

Nineteen patients with 19 sinus perforations were included in the study, 11 of whom had sinus perforations repaired with auto-transplanted teeth and 8 with flaps. The mean follow-up time for patients with AT and with FT was 24.82 months and 37.63 months, respectively. The mean age of patients with AT was 28.82, and the mean age of patients with FT was 34.36 (P > 0.05). More details are shown in Table 1.

**Table. 1.**

|  | Repair with AT | Repair with FT | P value |
| --- | --- | --- | --- |
| Number of patients | 11 | 8 | - |
| Number of sinuses | 11 | 8 | - |
| Age |  |  |  |
| Mean | 28.82 | 34.36 |  |
| Gender |  |  |  |
| Male | 6(54.5%) | 6(75%) |  |
| Female | 5(45.5%) | 2(25%) |  |
| Gingival index(GI) |  |  |  |
| 0 | 2 | 2 |  |
| 1 | 2 | 2 |  |
| 2 | 7 | 4 |  |
| Apical shadow<br>before surgery |  |  |  |
|  | 8(72.7%) | 7(87.5%) |  |
| Smoking |  |  |  |
| Yes | 2 | 3 |  |
| No | 9 | 5 |  |
| Follow-up<br>time(months) |  |  |  |
| Range | 20-35 | 16-53 |  |
| Mean | 24.82 | 37.63 |  |

The patients in the group who received AT underwent a follow-up visit two weeks after the operation. The soft tissue around the transplanted tooth exhibited slight swelling and loosening, categorized as I-II degrees. The root canal treatment was successfully completed and showed good healing. Subsequent follow-up examinations at six months and one year revealed further bone formation around the root of the donor tooth, with visible periodontal ligament images on the root apical radiograph. The maxillary sinus cavity appeared clean, and there was no thickening of the sinus mucosa.

The sutures were removed two weeks after the operation in the FT group. The wound had healed well at the one-month follow-up visit, with no signs of redness or mild swelling. CBCT scans taken three to six months later revealed a less continuous bone in the bottom wall of the maxillary sinus and a reduction in sinus mucosal thickness. The success rate of the AT group was 72.7% (8/11), and the success rate of the FT group was 100%. In the AT group, three patients exhibited widening of the periodontal ligament space and periapical low-density radiograph around their transplanted teeth. But there were no signs of loosening, redness, or swelling of the gums during the clinical examination. Additionally, the patients reported no discomfort, and therefore, the required follow-up was continued.

In 78.9%(15/19) of cases of maxillary sinus perforation, an apical shadow or thickening of the maxillary sinus mucosa was observed prior to tooth extraction. Among patients with maxillary sinus perforation, smokers accounted for 26.3% (5/19).

What’s more, the AT group was divided into three subgroups based on the gingival index (GI,0/1/2). The group with a GI score of 0 included 2 individuals, the group with a GI score of 1 included 2 individuals, and the group with a GI score of 2 included 7 individuals. The two-year survival probability of the AT group was assessed using Kaplan-Meier survival analysis. The analysis revealed that the survival probability was significantly lower when the GI score was 2 (51.8%) compared to when it was 0 or 1(100%)(Figure 5).

**Fig. 5.**
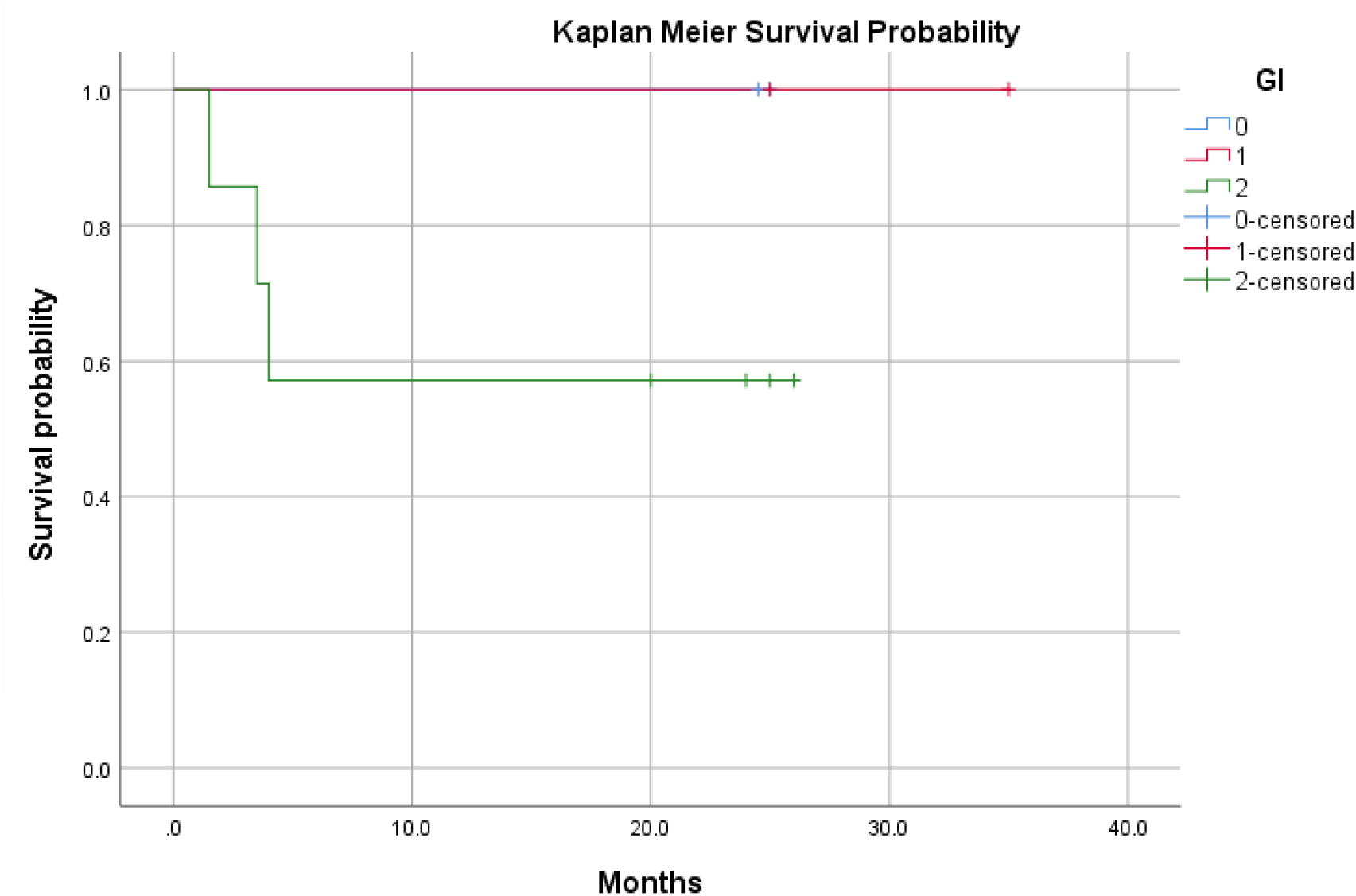

In comparison to the 72.7%(8/11) success rate of transplanted teeth, the flap group exhibited a higher success rate of 100%. All of the transplanted teeth were successfully retained, with good recovery of the gingival papilla and proper healing of the maxillary sinus perforation,the maxillary sinus cavity was clean, and the mucosa was not thickened; new bone formation was seen around the root of the donor tooth, and periodontal ligament images were seen on the root apical radiograph. The patients were able to perform chewing functions three months after the surgery. However, in the flap conversion group, an alternative repair method was required to restore masticatory function after three months. In this group, the gingival papilla disappeared, soft tissue scars were visible, the perforation of the maxillary sinus healed well.

## 4 Discussion

The purpose of this study is to compare the outcomes of auto-transplanted tooth restoration and flap restoration in repairing sinus perforation after tooth extraction. Additionally, it aims to discuss the feasibility of using auto-transplanted teeth combined with CGF for repairing maxillary sinus membrane perforation and restoring tooth extraction. Nineteen relevant cases were included in this study. The success rate of the postoperative AT group was 72.7% (8/11). All the donor teeth were preserved, the perforation of the maxillary sinus healed well, the cavity of the maxillary sinus was clean, and the mucosa was not thickened. The success rate of FT group was 100% (8/8), and the maxillary sinus perforation healed well. However, due to the small sample size of patients, more clinical cases and longer follow-up time are needed to verify the treatment plan. To the best of our knowledge, this is the first clinical study comparing the outcomes of auto-transplanted tooth and flap in repairing sinus perforation after tooth extraction.

When the maxillary sinus mucoperiosteal perforation exceeds 5 mm, the clinical treatment becomes more challenging[9]. Traditional repair methods for this condition include removable shield protection technology, surgical flap methods (such as buccal mucoperiosteal flap sliding, palatal mucoperiosteal flap transfer, and buccal fat pad transplantation), as well as autologous/non-autologous bone grafting techniques[10, 11]. However, these traditional surgical flap repair methods often disrupt the original gingival soft tissue morphology and structure, while autologous/non-autologous bone grafting techniques require effective sealing of the soft tissue to the graft[11, 13]. Furthermore, the traditional repair method necessitates postoperative implantation, causing additional trauma to the patient and preventing the restoration of the original gingival shape and structure[22]. In some cases, maxillary sinus lifting surgery may also be required, further complicating the implant restoration process. To address both maxillary sinus perforation and dentition defects simultaneously, the use of auto-transplanted teeth combined with CGF has shown promising results[23-25]. This approach allows for the retention of all transplanted teeth and the restoration of the gingival papilla, and maxillary sinus perforation healed well[26].

However, this surgical method has specific requirements for the surgical techniques used by the operator. It also necessitates postoperative root canal treatment by endodontic specialists[27, 28]. Additionally, the shape of the donor tooth must be compatible with the extraction socket of the recipient area[29]. Moreover, the mesial and distal areas of the edentulous area, as well as the vertical clearance, should not be too small[30].

The survival and success rates of auto-transplanted teeth are crucial indicators for the success of this procedure. Several studies have shown that the 1-year survival rate of transplanted teeth is 96%, while the 5-year survival rate is 84%[31]. The success rate is highest among young patients, reaching approximately 95%, and it is around 90% for patients under 30 years old and about 80% for patients over 30 years old[32, 33]. The activity and integrity of donor periodontal ligament cells play a vital role in the healing process of auto-transplanted teeth[33, 34]. The periodontal ligament (PDL) is a dense connective tissue that connects the cementum and alveolar bone[35]. It consists of fibers, matrix, and various types of cells, including periodontal ligament stem cells, cementoblasts, osteoblasts, and osteoclasts. These cells possess the ability to continuously proliferate and differentiate, actively contributing to the reconstruction and regeneration of alveolar bone and cementum[36, 37].

However, the study highlights the often overlooked aspect of patients’ oral hygiene status[38-40]. The findings indicate that when the gingival index (GI) is 2, the two-year survival probability of the AT group drops to 51.8%, which is significantly lower compared to when the GI is 0 or 1. Extensive research consistently emphasizes the importance of maintaining proper oral hygiene to prevent oral inflammation and promote the regeneration of periodontal tissues. Although the study had limitations in terms of the number of cases and follow-up time, it underscores the need to give sufficient attention to the oral hygiene status of postoperative patients.

Applying digital aided design in autologous tooth transplantation, along with the utilization of 3D printing models and guide plates, can lead to quicker and more effective alveolar socket preparation[19, 41]. This approach also helps in minimizing the need for trial planting, reducing operation time, minimizing damage to the periodontal ligament, and preserving periodontal ligament activity[42]. Consequently, it enhances the success rate of autologous tooth transplantation. Moreover, factors such as the stage of root development, postoperative root canal treatment, and the patient’s oral hygiene status also play crucial roles in determining the operation’s success[33, 43-45].

## 5 Conclusion

The subsequent results demonstrated that the utilization of auto-transplanted teeth had a positive impact on repairing maxillary sinus perforation. However, further studies with a larger sample size and longer follow-up duration are required to validate these findings. This procedure not only effectively seals the perforation of the maxillary sinus but also restores the dentition defect in the recipient area. Therefore, it appears to be a viable option for the restoration of patients with perforation of the maxillary sinus mucosa.

## Data Availability

All data produced in the present study are available upon reasonable request to the authors
All data produced in the present work are contained in the manuscript

## Notes

### Competing Interest Statement

The authors have declared no competing interest.

### Funding Statement

This study did not receive any funding

### Author Declarations

The study was approved by the Institutional Review Board of the Hospital of Stomatology Wuhan University

## References

1. Butaric, L.N., Differential Scaling Patterns in Maxillary Sinus Volume and Nasal Cavity Breadth Among Modern Humans. Anat Rec (Hoboken), 2015. 298(10): p. 1710–21.

2. Whyte, A. and R. Boeddinghaus, The maxillary sinus: physiology, development and imaging anatomy. Dentomaxillofac Radiol, 2019. 48(8): p. 20190205.

3. Rosano, G., et al., Maxillary sinus vascular anatomy and its relation to sinus lift surgery. Clin Oral Implants Res, 2011. 22(7): p. 711–715.

4. Güncü, G.N., et al., Location of posterior superior alveolar artery and evaluation of maxillary sinus anatomy with computerized tomography: a clinical study. Clin Oral Implants Res, 2011. 22(10): p. 1164–1167.

5. Marin, S., et al., Potential risk factors for maxillary sinus membrane perforation and treatment outcome analysis. Clin Implant Dent Relat Res, 2019. 21(1): p. 66–72.

6. Pommer, B., et al., Mechanical properties of the Schneiderian membrane in vitro. Clin Oral Implants Res, 2009. 20(6): p. 633–7.

7. Wang, X., et al., Association between smoking and Schneiderian membrane perforation during maxillary sinus floor augmentation: A systematic review and meta-analysis. Clin Implant Dent Relat Res, 2023. 25(1): p. 166–176.

8. Nemati, M., et al., Which factors affect the risk of membrane perforation in lateral window maxillary sinus elevation? A prospective cohort study. J Craniomaxillofac Surg, 2023.

9. Cui, Q.Y., et al., [A preliminary exploration into the efficacy of personalized surgical schemes in the repair of maxillary sinus perforation and maxillary sinus fistula]. Zhonghua Kou Qiang Yi Xue Za Zhi, 2022. 57(9): p. 953–957.

10. Yalçın, S., et al., Surgical treatment of oroantral fistulas: a clinical study of 23 cases. J Oral Maxillofac Surg, 2011. 69(2): p. 333–9.

11. Alkan, A., et al., The reconstruction of oral defects with buccal fat pad. Swiss Med Wkly, 2003. 133(33-34): p. 465–70.

12. Scott, P., G. Fabbroni, and D.A. Mitchell, The buccal fat pad in the closure of oro-antral communications: an illustrated guide. Dent Update, 2004. 31(6): p. 363–4, 366.

13. Abuabara, A., et al., Evaluation of different treatments for oroantral/oronasal communications: experience of 112 cases. Int J Oral Maxillofac Surg, 2006. 35(2): p. 155–8.

14. Al-Moraissi, E., et al., Does intraoperative perforation of Schneiderian membrane during sinus lift surgery causes an increased the risk of implants failure?: A systematic review and meta regression analysis. Clin Implant Dent Relat Res, 2018. 20(5): p. 882–889.

15. Park, W.B., K.L. Kang, and J.Y. Han, Factors influencing long-term survival rates of implants placed simultaneously with lateral maxillary sinus floor augmentation: A 6-to 20-year retrospective study. Clin Oral Implants Res, 2019. 30(10): p. 977–988.

16. Ashurko, I., et al., Delayed autotransplantation as a method of single defect treatment with sinus perforation. Case report. J Clin Exp Dent, 2023. 15(2): p. e160–e164.

17. Kitagawa, Y., et al., Use of third molar transplantation for closure of the oroantral communication after tooth extraction: a report of 2 cases. Oral Surg Oral Med Oral Pathol Oral Radiol Endod, 2003. 95(4): p. 409–15.

18. Verweij, J.P., et al., Replacing Heavily Damaged Teeth by Third Molar Autotransplantation With the Use of Cone-Beam Computed Tomography and Rapid Prototyping. J Oral Maxillofac Surg, 2017. 75(9): p. 1809–1816.

19. Ashkenazi, M., et al., Computerized three-dimensional design for accurate orienting and dimensioning artificial dental socket for tooth autotransplantation. Quintessence Int, 2018. 49(8): p. 663–671.

20. Assis, R.I.F., et al., Osteogenic Commitment of Human Periodontal Ligament Cells Is Predetermined by Methylation, Chromatin Accessibility and Expression of Key Transcription Factors. Cells, 2022. 11(7).

21. Wang, J., et al., Effects of platelet-rich fibrin on osteogenic differentiation of Schneiderian membrane derived mesenchymal stem cells and bone formation in maxillary sinus. Cell Commun Signal, 2022. 20(1): p. 88.

22. de Freitas Coutinho, N.B., et al., Success, Survival Rate, and Soft Tissue Esthetic of Tooth Autotransplantation. J Endod, 2021. 47(3): p. 391–396.

23. Koleilat, A., et al., A Combination of Platelet-Rich Fibrin and Collagen Membranes for Sinus Membrane Repair: A Case Report (Repair of Sinus Membrane Perforation). Dent J (Basel), 2023. 11(3).

24. Dong, K., et al., The extract of concentrated growth factor enhances osteogenic activity of osteoblast through PI3K/AKT pathway and promotes bone regeneration in vivo. Int J Implant Dent, 2021. 7(1): p. 70.

25. Gonzalez-Ocasio, J. and M. Stevens, Autotransplantation of Third Molars With Platelet-Rich Plasma for Immediate Replacement of Extracted Non-Restorable Teeth: A Case Series. J Oral Maxillofac Surg, 2017. 75(9): p. 1833.e1-1833.e6.

26. Melillo, M. and L. Boschini, Autotransplantation of third molar for oro-antral communication closure: evidence of sinus elevation after healing. Minerva Dent Oral Sci, 2021. 70(4): p. 169–171.

27. Hwang, L.A., et al., Rapid prototyping-assisted tooth autotransplantation is associated with a reduced root canal treatment rate: a retrospective cohort study. BMC Oral Health, 2022. 22(1): p. 25.

28. Lin, P.Y., et al., Endodontic considerations of survival rate for autotransplanted third molars: a nationwide population-based study. Int Endod J, 2020. 53(6): p. 733–741.

29. Ong, D., Y. Itskovich, and G. Dance, Autotransplantation: a viable treatment option for adolescent patients with significantly compromised teeth. Aust Dent J, 2016. 61(4): p. 396–407.

30. Martin, K., S. Nathwani, and R. Bunyan, Autotransplantation of teeth: an evidence-based approach. Br Dent J, 2018. 224(11): p. 861–864.

31. Sugai, T., et al., Clinical study on prognostic factors for autotransplantation of teeth with complete root formation. Int J Oral Maxillofac Surg, 2010. 39(12): p. 1193–203.

32. Tsukiboshi, M., N. Yamauchi, and Y. Tsukiboshi, Long-term outcomes of autotransplantation of teeth: A case series. Dent Traumatol, 2019. 35(6): p. 358–367.

33. Kafourou, V., et al., Outcomes and prognostic factors that influence the success of tooth autotransplantation in children and adolescents. Dent Traumatol, 2017. 33(5): p. 393–399.

34. Liu, J., et al., Macrophage polarization in periodontal ligament stem cells enhanced periodontal regeneration. Stem Cell Res Ther, 2019. 10(1): p. 320.

35. Connizzo, B.K., et al., Nonuniformity in Periodontal Ligament: Mechanics and Matrix Composition. J Dent Res, 2021. 100(2): p. 179–186.

36. Xu, J., et al., Periodontal Ligament Stem Cell-Derived Extracellular Vesicles Enhance Tension-Induced Osteogenesis. ACS Biomater Sci Eng, 2023. 9(1): p. 388–398.

37. Huang, H.M., et al., Mechanical force-promoted osteoclastic differentiation via periodontal ligament stem cell exosomal protein ANXA3. Stem Cell Reports, 2022. 17(8): p. 1842–1858.

38. Luo, H., et al., Oral Health, Diabetes, and Inflammation: Effects of Oral Hygiene Behaviour. Int Dent J, 2022. 72(4): p. 484–490.

39. Preus, H.R., Q. Al-Lami, and V. Baelum, Oral hygiene revisited. The clinical effect of a prolonged oral hygiene phase prior to periodontal therapy in periodontitis patients. A randomized clinical study. J Clin Periodontol, 2020. 47(1): p. 36–42.

40. Reynolds, M.A., et al., Periodontal regeneration - intrabony defects: a consensus report from the AAP Regeneration Workshop. J Periodontol, 2015. 86(2 Suppl): p. S105–7.

41. Abella, F., et al., Outcome of Autotransplantation of Mature Third Molars Using 3-dimensional-printed Guiding Templates and Donor Tooth Replicas. J Endod, 2018. 44(10): p. 1567–1574.

42. Chai, H.Y., et al., Use of three-dimensional-printed tooth replica for autogenous tooth transplantation at an anatomically challenging site. J Dent Sci, 2023. 18(1): p. 472–474.

43. Jang, Y., et al., Prognostic Factors for Clinical Outcomes in Autotransplantation of Teeth with Complete Root Formation: Survival Analysis for up to 12 Years. J Endod, 2016. 42(2): p. 198–205.

44. Almpani, K., S.N. Papageorgiou, and M.A. Papadopoulos, Autotransplantation of teeth in humans: a systematic review and meta-analysis. Clin Oral Investig, 2015. 19(6): p. 1157–79.

45. Yang, S., B.Y. Jung, and N.S. Pang, Outcomes of autotransplanted teeth and prognostic factors: a 10-year retrospective study. Clin Oral Investig, 2019. 23(1): p. 87–98.

